# Nationwide Seroprevalence of SARS-CoV-2 in Saudi Arabia

**DOI:** 10.1101/2021.01.28.21250598

**Authors:** Naif Khalaf Alharbi, Suliman Alghnam, Abdullah Algaissi, Hind Albalawi, Mohammed W. Alenazi, Areeb M. Albargawi, Abdullah G. Alharbi, Abdulaziz Alhazmi, Ali Al Qarni, Ali Alfarhan, Hossam M. Zowawi, Hind Alhatmi, Jahad Alghamdi, Fayhan Alroqi, Yaseen M. Arabi, Anwar M. Hashem, Mohammed Bosaeed, Omar Aldibasi

## Abstract

**Background:** Estimated seroprevalence of Coronavirus Infectious Disease 2019 (COVID-19), caused by the Severe Acute Respiratory Syndrome-Coronavirus-2 (SARS-CoV-2) is a critical evidence for a better evaluation of the virus spread and monitoring the progress of the COVID-19 pandemic in a population. In the Kingdom of Saudi Arabia (KSA), SARS-CoV-2 seroprevalence has been reported in specific regions, but an extensive nationwide study has not been reported. Here, we report a nationwide study to determine the prevalence of SARS-CoV-2 in the population of KSA during the pandemic, using serum samples from healthy blood donors, non-COVID patients and healthcare workers (HCWs) in six different regions of the kingdom, with addition samples from COVID-19 patients.

**Methods:** A total of 11703 serum samples were collected from different regions of the KSA including; 5395 samples from residual healthy blood donors (D); 5877 samples from non-COVID patients collected through residual sera at clinical biochemistry labs from non-COVID patients (P); and 400 samples from consented HCWs. To determine the seroprevalence of SARS-CoV-2, all serum samples, in addition to positive control sera from RT-PCR confirmed COVID-19 patients, were subjected to in-house ELISA with a sample pooling strategy, which was further validated by testing individual samples that make up some of the pools, with a statistical estimation method to report seroprevalence estimates

**Results:** Overall (combining D and P groups) seroprevalence estimate was around 11% in Saudi Arabia; and was 5.1% (Riyadh), 1.5% (Jazan), 18.4% (Qassim), 20.8% (Hail), 14.7% (ER; Alahsa), and 18.8% in Makkah. Makkah samples were only D group and had a rate of 24.4% and 12.8% in the cities of Makkah and Jeddah, respectively. The seroprevalence in Saudi Arabia across the sampled areas would be 12 times the COVID-19 infection rate. Among HCWs, 7.5% (4.95-10.16 CI 95%) had reactive antibodies to SARS-CoV-2 without reporting any previously confirmed infection. This was higher in HCWs with hypertension. The study also presents the demographics and prevalence of co-morbidities in HCWs and subset of non-COVID-19 population.

**Conclusion:** Our study estimates the overall national serological prevalence of COVID-19 in Saudi Arabia to be 11%, with an apparent disparity between regions.

## Introduction

The severe acute respiratory syndrome coronavirus-2 (SARS-CoV-2) is a novel coronavirus that was first identified in Wuhan city in China by the end of 2019 and spread globally to cause epidemics in almost all parts of the world. In March 2020, the World Health Organization declared it a global pandemic. The disease, named coronavirus disease 2019 (COVID-19), causes a respiratory illness that could range from severe pneumonia to mild respiratory illness with symptoms such as fever, dry cough, fatigue, headaches, shortness of breath and gastroenteritis (diarrhea). However, some cases are completely asymptomatic (1). As of January 2021, there have been more than 90 million confirmed cases with ∼1.9 million deaths worldwide (1).

While immune responses against SARS-CoV-2 could be induced as early as the first week after symptoms onset, seroconversion usually occurs at a median of 10-12 days for IgM and 12-15 days for IgG and could reach 100% by 21 days in most infected individuals (2,3,4). Serum IgM levels peak at two to three weeks, whereas the IgG antibodies peak at three to four weeks post symptoms (4). This increase of antibodies over time is usually accompanied by decreased viral RNA in the respiratory tract (3). The neutralizing activity of these antibodies, particularly IgG, peaks at one month and could last for up to eight months post symptoms onset (5). Nonetheless, some reports suggest a decline in the levels of neutralizing antibodies (NAb) after three months post-infection (6).

The severity of cases is usually associated with earlier antibody induction and higher levels of such antibodies (2). Therefore, asymptomatic cases may only show low levels of antibodies that would significantly decline in a short time. It has been observed that 40% of asymptomatic cases would become seronegative in two months (7–9), which may post challenges to assess the seroprevalence of COVID-19 in the general healthy population especially that seroprevalence is an indicator of asymptomatic or unreported cases. While most symptomatic COVID-19 cases would usually be captured and reported clinically and by diagnostic means, seroprevalence studies would mainly reveal the asymptomatic or very mild cases.

Seroprevalence studies on COVID-19 are being conducted in many countries to estimate the true prevalence of COVID-19. These studies aid in defining the disease burden, gauging the need for vaccine coverage, establishing the correlates of protection, evaluating the possibility and impact of re-infections, and evaluating the benefits of public health measures such as lockdown and travel bans. Studies showed that the seroprevalence of COVID-19 in general populations varies across the world. For example, it ranges between 17% in Iran (10) to 5% in Spain (11), with limited studies that evaluated seroprevalence at national levels. According to the SeroTracker database, there have been more than 300 serological studies worldwide; estimating the seroprevalence to be 2.14% in blood donors, 3.22% in Healthcare Workers (HCWs), and 6.48% in general populations as per several estimates from different countries worldwide, with a wide range of 95% CI (12). Another metaanalysis has reported that the seroprevalence in general population ranges from 0.37% to 22.1% (13). However, there is still a need for conducting more seroprevalence studies to provide information considering the variability of specific populations, time of sampling, locations, and type of the measured immune responses.

There is a need to study and report seroprevalence rates in various populations in the Middle East and Kingdom of Saudi Arabia (KSA); especially that KSA implemented an early nationwide lockdown, banned domestic and international travels by any transportation, closed all places of leisure, shopping, utilities, and worship. There were only two large-sample serological studies reported from KSA: one focused on the magnitude and longevity of antibodies in recovered COVID-19 patients (8) and the second on seropositivity rate in HCWs across the country, which reported a national seroprevalence rate of 2.36% in HCWs (14). However, an extensive nationwide study has not been conducted.

Therefore, we here present a nationwide COVID-19 serological prevalence study in KSA with serum samples collected from four different populations in six provinces of the country. The populations are healthy blood donors (D), non-COVID-19 patients (P), confirmed COVID-19 patients, and HCWs. The sampled regions are the provinces of Riyadh (RYD), Jazan (JZN), Qassim (QSM), Hail, Eastern Region (ER – Alahsa governorate area), and Makkah. The latter was divided into Makkah City (MKH) and Jeddah City (JDH) in order to highlight the burden of COVID-19 and seroprevalence in the holy city of Makkah since it is a travel destination for millions of pilgrims from across the globe and would support more efficient global health measures. This study present estimated seroprevalence based on an in-house ELISA with a sample pooling strategy, which was further validated by testing individual samples that make up some of the pools, with a statistical estimation method. This study also presents the chronic diseases and demographics of a subset of non-COVID-19 patients and HCWs.

## Materials and Methods

### Samples and subjects

A total of 11703 serum samples were collected from different regions including 5395 residual samples from healthy blood donors (D); 5877 samples from residual sera at clinical biochemistry labs from non-COVID patients (P); 31 samples from consented infected COVID-19 patients within ten days of infection; and 400 samples from consented HCWs.

Samples collected from D and P were from six Saudi Arabian provinces (regions): Riyadh (RYD), Jazan (JZN), Qassim (QSM), Hail, Eastern Region (ER, Alahsa governorate in particular), and Makkah Region in which samples were collected from the cities of Makkah (MKH) and Jeddah (JDH). From the following hospitals: National Guard Health Affairs (NGHA) hospitals in Riyadh and Alahsa, Jazan University Hospital in Jazan, Central Blood Bank in Qassim, Central Blood Bank in Hail, King Abdullah Medical Complex in Jeddah and Central Blood Bank in Makkah. Samples from HCWs and COVID-19 patients were collected from Prince Mohammad bin Abdulaziz hospital and NGHA hospital in Riyadh city. The non-COVID-19 patients were defined as any person who is visiting a clinic or admitted at a hospital with samples collected for clinical biochemistry blood testing and confirmed to be negative for COVID-19 by RT-PCR.

### ELISA

An in-house enzyme-linked immunosorbent assay (ELISA) was developed and applied to detect IgG against SARS-CoV-2 in serum samples. Nunc MaxiSorp 96-well ELISA microplates (Thermo Fisher, Waltham, MA) were coated with recombinant S1 subunit of the SARS-CoV-2 Spike protein (Sino Biological, China) at a concentration of 1 μg/ml. The plates were incubated overnight at room temperature (RT). Plates were then washed six times with washing buffer (phosphate buffered saline (PBS) with 0.5% Tween20, PBS-T) using automated Microplate Washer (Molecular Devices, San Jose, CA). Then, wells were blocked by washing buffer containing 10% skimmed milk (blocking buffer) for one hour at RT. Serum samples from COVID-19 patients and HCWs were diluted at 1:100 in PBS-T, and 50 μl of each diluted sample were added into duplicate wells and incubated for two hours. Samples from blood donors and non-covid-19 patient were prepared in pools of 10 samples by adding 2 μl from each serum sample to 180 μl of PBS-T (i.e. 1:100 dilution), and 50 μl of each diluted pool were added into duplicate wells and incubated for two hours. Some positive pooled samples were subsequently tested as individual samples for confirmation. Plates were washed, and 50 μl of 1:1000 diluted alkaline phosphatase labeled goat anti-human IgG secondary antibody (Thermo Fisher, Waltham, MA) were added and incubated for one hour at RT. After washing, a substrate made of PNPP (pnitrophenylphosphate, sigma) tablets dissolved in diethanolamine buffer solution and distal water was added. This was followed by measuring optical density (OD) at 405 nm using Microplate Reader (Molecular Devices, San Jose, CA). The cut-off value was set as the average of the negative control serum samples plus three times of standard deviation. Negative control samples were sera collected before the COVID-19 pandemic, and positive control samples were from confirmed recovered COVID-19 cases. The same control samples were aliquoted and used in every ELISA run. ELISA was repeated on different pools from all regions in order to confirm positivity.

### Confirmatory ELISA

Two confirmatory ELISA tests were performed in order to confirm the pooled sample results and assess the average number of positive samples per pool. First, individual samples from 19 pools (n=190) were evaluated using anti-SARS-CoV-2 ELISA IgA (EUROIMMUN, Germany) following the manufacturer’s instruction. Second, individual samples from 57 pools (n=570), ten pools from each region (5 from each group; donors and non-COVID-19 patients) were evaluated using the in-house anti-SARS-CoV-2 spike IgG ELISA as described above.

### Statistical Analysis

Samples pooling strategy has been recommended to expand test coverage and save resources by eliminating the number of tests required for diagnostic, screening, and surveillance programs (15,16). Our estimates are derived based on 10 samples per pool. Thus, a positive pool includes at least one (10%) positive sample. Stratified by region and population, we obtained the average number of positive samples in positive pools. Then, we randomly selected five positive pools per population from all regions, except for Jazan D population, for individual testing to obtain the average number of positive individual samples per population per region. Therefore, the prevalence for each region and population were obtained based on the equation below with 95% confidence intervals obtained using the pool’s standard error of the mean.

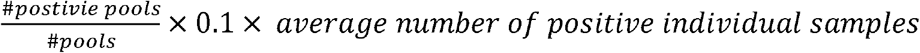

Demographic information were summarized using descriptive statistics in terms of mean and standard deviation or frequencies and percentages. For the chronic diseases in the HCWs in Riyadh, group comparisons were made based on chi-square tests, and the level of significance is alpha = 0.5. All statistical analyses were conducted using SAS 9.4 (SAS Institute Inc., Cary, NC) and GraphPad Prism V8 software (GraphPad Co.).

### Ethical Approval

The study was approved by the IRB in KAIMRC (Ministry of National Guard Health Affairs) for project number RC20-180. Residual samples from blood banks and clinical laboratories were obtained after an IRB approval. COVID-19 patients and HCWs signed informed consents to give blood samples.

## Results

### Demographics and chronic diseases of the study subjects

The blood donor population (group D) was predominantly male with an average age ranging from 26 to 32 in the sampled regions; the non-COVID-19 patient population (group P) was balanced in terms of gender distribution in most regions with an age range of 24 to 58 in different regions (Table S1). Among the P group (n=5877), 3,592 subjects sampled in Eastern Region (Alahsa) and Riyadh region were further analyzed to highlight the presence of chronic diseases, which includes hypertension, diabetes, asthma, and dyslipidemia. The prevalence of chronic diseases in this subset population of non-COVID-19 patients were 51.7% obesity (BMI >40), 45% diabetes, 49% hyperlipidemia, 35% hypertension, and 3.3% asthma. No formal association was done with the seroprevalence due to the nature of pooling sera for testing.

### National seroprevalence of SARS-CoV-2 in Saudi Arabia

To estimate the COVID-19 seroprevalence in KSA, a total of 11,272 samples were collected from six provinces of KSA; 5395 samples were collected from the blood donors and 5877 samples from the non-COVID-19 patients (Table 1). These sera were tested in pools of 10 samples, and the results showed high seropositivity in pooled samples compared to negative and positive quality control samples (Figure 1 A and B). The positive control samples were collected from two confirmed COVID-19 recovered cases and the negative control samples were collected prior to COVID-19 pandemic. Individual serum samples from 57 selected positive pools (10 pools per region; 5 from group D and 5 from group P, except 2 pools from Jazan blood donors since they were negative) were tested for anti-SARS-CoV-2 spike IgG (Figure 1C). Individual samples were selected from pools that have a range of higher and lower OD values, although OD value is not quantitative. Further, individual serum samples from 19 selected positive pools were tested for anti-spike IgA ELISA (Figure 1D).

**Table 1:** Estimated seroprevalence rate of COVID-19 in six Saudi Arabian regions based on blood donors and non-COVID-19 patients

| Region | Population size | COVID-19 incident rate (%) | No of samples | Sample collection time | Seroprevalence (%) | Seroprevalence 95% CI |
| --- | --- | --- | --- | --- | --- | --- |
| **Sampled regions** | **24,750,893** | **1.06** | **11275** | **Jun-Nov 2020** | **10.9** | **(10.3,11.6)** |
| Blood Donors |  |  | 5385 |  | 8.8 | (8,9.7) |
| Non-COVID-19 Patients |  |  | 5890 |  | 13.3 | (12.4,14.2) |
| **Riyadh** | **8,014,678** | **0.881** | **4237** | **20-Jun** | **5.1** | **(4.5,0.8)** |
| Blood Donors |  |  | 1490 |  | 2.4 | (1.5,3.2) |
| Non-COVID-19 Patients |  |  | 2747 |  | 6.3 | (5.5,0.1) |
| **Jazan** | **1,535,167** | **0.78** | **640** | **20-Jun** | **1.6** | **(0.7,2.5)** |
| Blood Donors |  |  | 80 |  | 0 | (0,0) |
| Non-COVID-19 Patients |  |  | 560 |  | 1.8 | (0.8,2.8) |
| **Qassim** | **1,389,929** | **0.92** | **3032** | **Jul-Aug 2020** | **18.4** | **(17,19.8)** |
| Blood Donors |  |  | 1632 |  | 13.7 | (11.8,15.6) |
| Non-COVID-19 Patients |  |  | 1400 |  | 24.5 | (22.8,26.2) |
| **Hail** | **685,423** | **0.98** | **460** | **Jul-Aug 2020** | **20.9** | **(18.3,23.5)** |
| Blood Donors |  |  | 300 |  | 8 | (6.5,9.5) |
| Non-COVID-19 Patients |  |  | 160 |  | 42.2 | (36.7,47.7) |
| **Alahsa** | **4,787,375** | **1.8** | **1874** | **20-Nov** | **14.7** | **(12.7,16.6)** |
| Blood Donors |  |  | 851 |  | 10.6 | (7.6,13.6) |
| Non-COVID-19 Patients |  |  | 1023 |  | 16.8 | (14.7,19) |
| **Makkah** | **8,338,321** | **1** | **1032** | **20-Nov** | **18.8** | **(15.8,21.9)** |
| Blood Donors Makkah City |  |  | 532 |  | 24.5 | (22.3,26.6) |
| Blood Donors Jeddah City |  |  | 500 |  | 12.9 | (9,16.8) |

**Figure 1:**
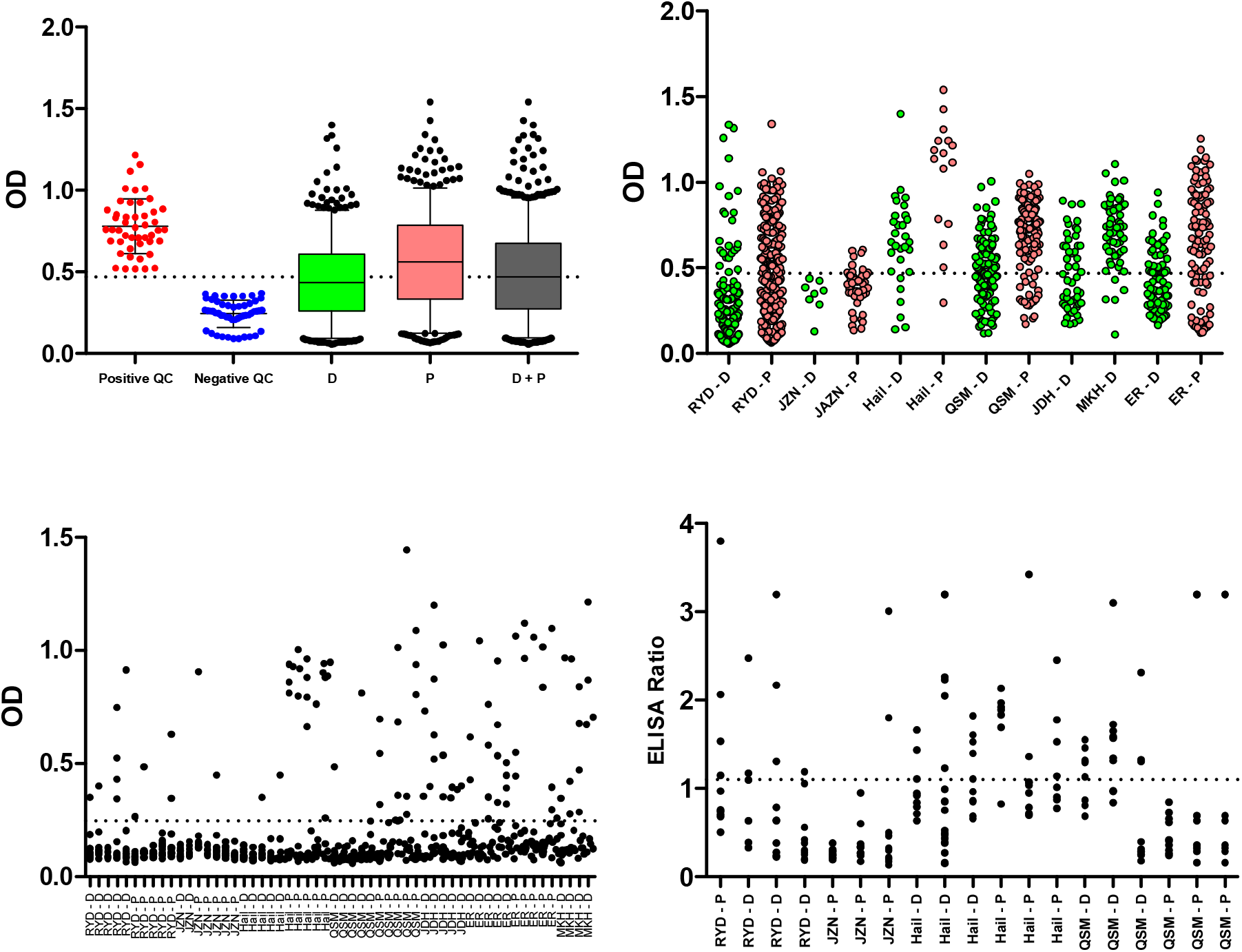
Anti-SARS-CoV-2 antibodies in serum samples from Saudi Arabia. In-house ELISA for anti-SARS-CoV-2 spike IgG antibodies (A, B, and C). **A:** Serum samples from blood donors (D; green) and non-COVID-19 patients (P; red) were pooled (10 samples per pool) and evaluated along with positive and negative quality control (QC) samples. The same control samples were tested in quadruplicates in each ELISA plate and values from different plates are plotted. **B:** Pooled samples (10 per pool) from D (green) and P (red) are plotted for each region. Regions’ names were included as described in the text. Individual samples that made up some of the pools were further tested using in-house anti-spike IgG ELISA (**C)** or using commercial anti-spike IgA ELISA (**D**). The in-house ELISA’s cut-off value was calculated as the average of negative controls plus three times the standard deviation. The cut-off value for commercial ELISA was 1.1. QC: Quality control samples.

The average number of positive individual samples per pool was used to calculate the seroprevalence per population and per region. Regional seroprevalence in the D group was 2.36% in Riyadh, 0% in Jazan, 13.65% in Qassim, 8% in Hail, 10.62 in Eastern Region (Alahsa areas), and 18.84% in Makkah (24.45% and 12.88 in the cities of Makkah and Jeddah, respectively). Regional seroprevalence in the P group was 6.30% in Riyadh, 1.78% in Jazan, 24.49% in Qassim, 42.18% in Hail, and 16.84 in Eastern Region (Alahsa areas). There was a trend of higher seroprevalence estimate in P group as compared to D group (Figure 1A). The overall regional seroprevalence estimate (combining D and P groups) was 11% in Saudi Arabia; and was 5% in Riyadh, 1.5% in Jazan, 18% in Qassim, 20.8% in Hail, and 14.7% in Alahsa. It is important to note that the sample collection time was from Jun to Aug, except for Alahsa, Makkah, and Jeddah sampling which was done through November (Table 1). The seroprevalence estimates were compared to the COVID-19 incident (infection) rate in December 2020, in the sampled regions, showing a trend of increasing seroprevalence in regions that have higher COVID-19 incident rate (Sampled regions in Table 1 and Figure 2; unsampled regions in Table S2). This indicates that the seroprevalence rate is more than the incident rate by around 6x (Riyadh), 2x (Jazan), 20x (Qassim), 21x (Hail), 8x (Alahsa), and 18x (Makkah), were x means how many times. Based on this, the seroprevalence in Saudi Arabia across the sampled regions would be 12x more than the incident rate (Table 1 and Figure 2).

**Figure 2:**
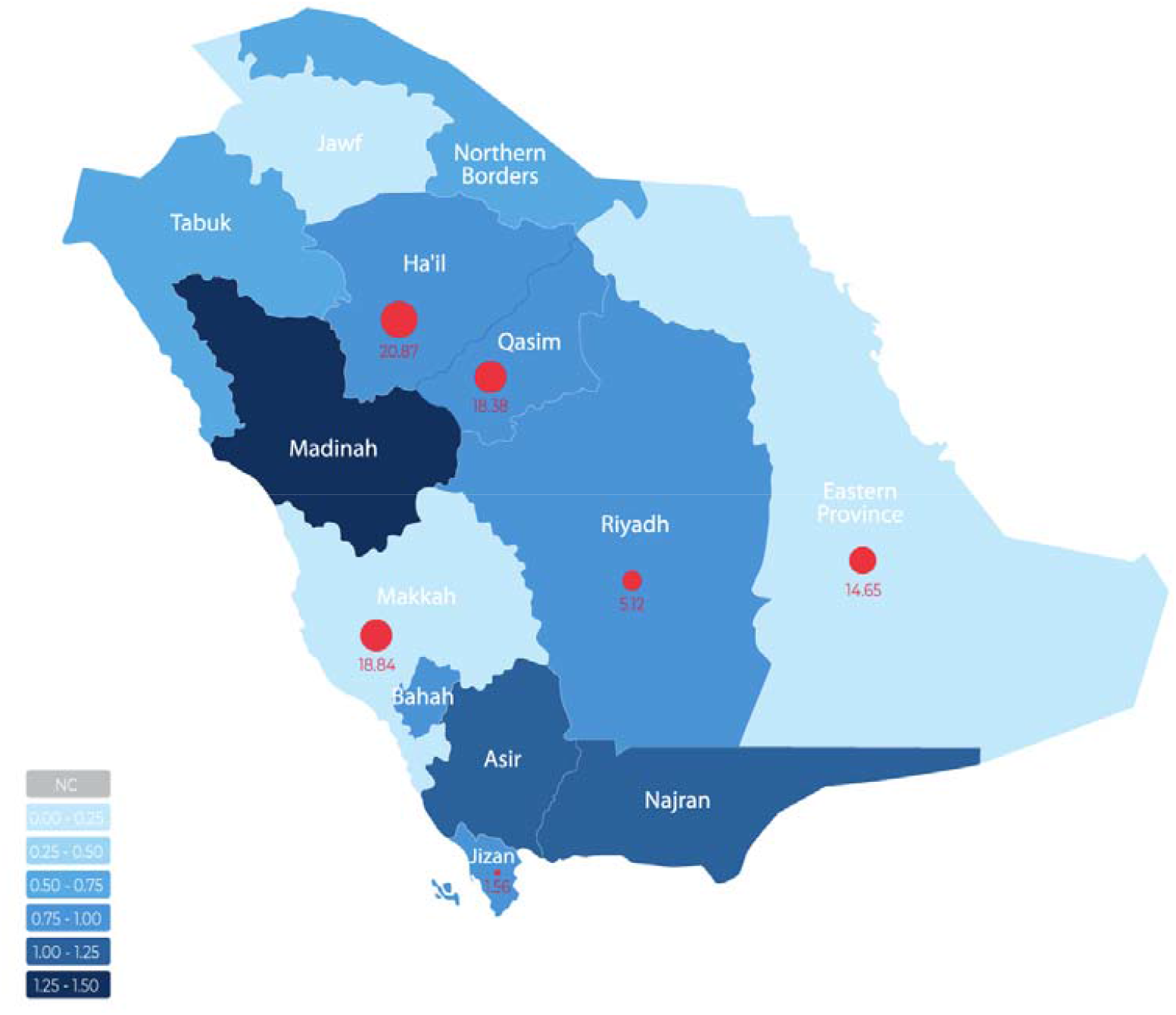
Estimated seroprevalence rate of COVID-19 and COVID-19 infection rates in all sampled and unsampled Saudi Arabian regions. Geographical map showing the 13 regions (provinces) of Saudi Arabia. The blue colour intensity represents the COVID-19 incident rates according to the Saudi Ministry of Health; as shown in Table S2. Red circles represent seroprevalence estimates with a circle size relevant to estimates.

### Seroprevalence of SARS-CoV-2 in Healthcare Workers in Riyadh, Saudi Arabia

To estimate the COVID-19 seroprevalence in HCWs in Riyadh city, a total of 400 samples were collected from health practitioners (doctors, nurses, and workers in hospital wards) from two of the main COVID-19 hospitals in Riyadh city; where high number of COVID-19 patients were admitted and treated. Among these HCWs, 7.5% (4.95-10.16 CI 95%) had reactive antibodies to SARS-CoV-2 without reporting any previously confirmed infection (Figure 3A). Most of HCWs (n=294) gave access to their clinical and demographic information (Table 2). There was a significant relation between seropositive HCWs and hypertension but no association with any of the remaining co-morbidities such as diabetes and obesity was observed. In addition, we included serum samples from 31 acute COVID-19 cases, from the same hospitals, that showed the acceptability of the in-house anti-SARS-CoV-2 spike IgG (Figure 3B).

**Table 2:** Demographics and chronic diseases in healthcare workers in Riyadh, Saudi Arabia.

|  | Seronegative | Seropositive | P value |
| --- | --- | --- | --- |
| Average age | 40 | 37.1 | 0.3 |
| Male | N (%)<br>107 (39.4) | N (%)<br>6 (31.5) | 0.49 |
| Diabetes | N (%)<br>36 (13.2) | N (%)<br>2 (10.5) | 0.73 |
| Hypertension | N (%)<br>57 (20.9) | N (%)<br>10 (52.6) | **0.02** |
| Asthma | N (%)<br>1 (0.37) | N (%)<br>1 (5.2) | 0.12 |
| Obesity | N (%)<br>44 (19.3) | N (%)<br>2 (12.5) | 0.29 |
| Depression | N (%)<br>5 (1.8) | N (%)<br>1 (5.2) | 0.33 |
| Hyperlipidemia | N (%)<br>58(21.3) | N (%)<br>5 (26.3) | 0.33 |

**Figure 3:**
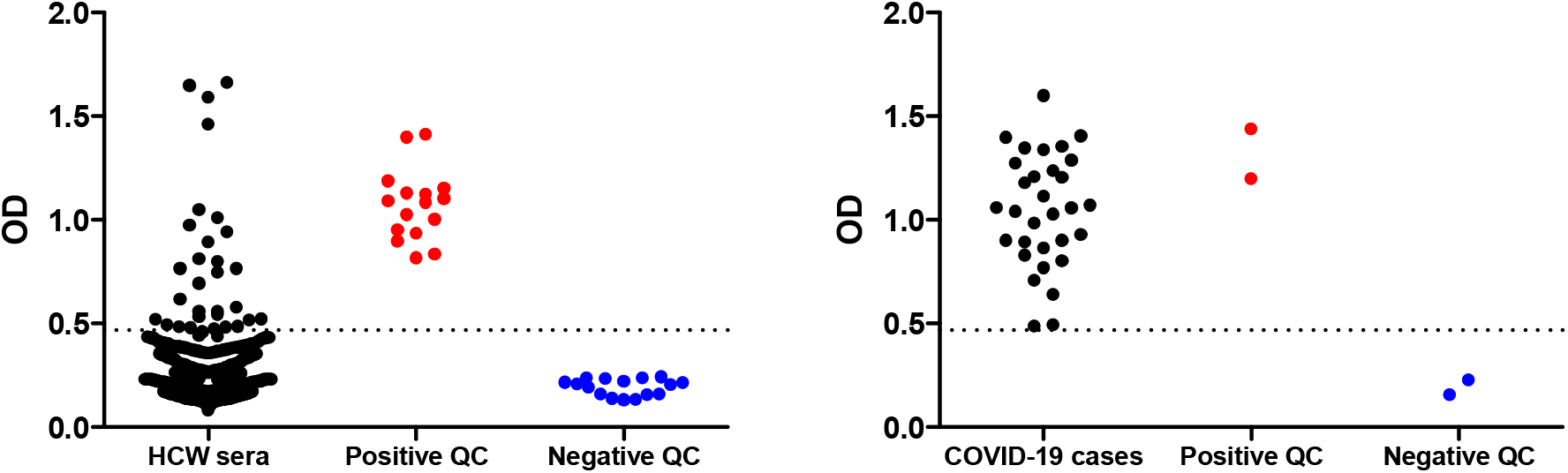
Anti-SARS-CoV-2 antibodies in samples from COVID-19 patients and HCWs in Riyadh, Saudi Arabia. In-house ELISA for anti-SARS-CoV-2 spike IgG antibodies in individual serum samples from HCWs (**A**) and acute COVID-19 cases (**B**) with positive and negative controls included as in figure 1. The in-house ELISA’s cut-off value was calculated as the average of negative controls plus three times the standard deviation.

## Discussion

In this study, serum samples were collected from over 11,000 persons in order to estimate the national seroprevalence of COVID-19 in Saudi Arabia; samples were collected from six (out of 13) geographically distributed regions of the country. These regions have a total population of 24.75 million, a majority out of the 34.8 million total Saudi population. A sample pooling strategy was applied to detect anti-SARS-CoV-2 spike IgG antibodies using an in-house ELISA; followed by testing individual samples from some pools to estimate the seroprevalence. The individual sample testing was also performed using different ELISAs that detects anti-SARS-CoV-2 IgA and IgG. The collected samples were mainly from healthy blood donors and residual samples from diagnostic laboratories, collected from in- and out-patients (non-COVID-19 patients).

This approach has estimated the overall seroprevalence of COVID-19 in Saudi Arabia to be 11%, ranging from 1.78 to 24.45% in different regions. The seroprevalence among blood donors, and non-COVID patients and HCWs are substantial, suggesting high rates of asymptomatic or undiagnosed COVID-19. Non-COVID patients had a trend of higher seroprevalence than blood donors, likely related to co-morbid conditions or indication for hospital stays or visits. It is important to note that the time of sample collection varied across regions; however, that did not have a major effect since the seroprevalence is shown to be increasing with the incident rate of COVID-19 in December 2020. For example, Jazan region was sampled early in June and had the lowest seroprevalence, but it has a low incident rate as of December 2020. In unsampled regions, the seroprevalence would vary according to the disease incident. For example, Madinah region had the highest incident rate in KSA (1.27%), Table S2, and was not sampled in our study, but a recent study has found the local seroprevalence in Madinah to be 19.3% in blood donors (18), falling in a similar range to Makkah regions that had 1% incident rate and 18.8% seroprevalence estimate. However, time of serological study is crucial, a study has reported a seroprevalence of 0% in blood donor samples (n=956) in Jeddah, collected between January and May 2020 (19). Of note, Saudi Arabia has detected its first COVID-19 case on the 2^nd^ of March 2020; and by 12^th^ of January 2021, the incident rate in the country was around 1% (363,259 confirmed cases in the Saudi population of 34.8 million) (17).

A number of studies have reported seroprevalence in blood donors, identifying the rate to be 34.2% in Pakistan, 25% in India, 7.7% in Brazil, and 6.5% in the UK; however, only a few studies have reported at a national scale. A total of 14 studies in the UK have identified that the seroprevalence rate is 14%. For residual samples (resembling our non-COVID-19 patient population), studies have identified a rate of 3.97% in the UK and 10.72% in France (SeroTracker.com). SeroTracker.com shows that seroprevalence studies can be very different in terms of serological assays, sample source, geographical coverage, and the population type; in addition, the timing of these studies may only reflect the dynamic transmission of the virus at the time. It may not be practical to compare our data with any other international seroprevalence reports. In Saudi Arabia, there was no previous national report on the general public, blood donors, or residual samples. Studies on HCWs in Saudi Arabia showed that the seroprevalence is 24% in in Madinah city (20) and 2.36% nationally (14). Our data on HCWs sampled from two of the COVID-19 leading hospitals in the Capital Riyadh showed a rate of 7.5%. Considering age, gender, and co-morbidities in our HCWs, there was a significant indication of an association between seroprevalence in HCWs and prevalence of hypertension, although this is a small size cohort and requires further studies on a larger sample size.

This study has strengths of sampling largely populated, geographically distributed regions; but it has its limitation of using proxy samples and it had a pooling strategy that may not reflect precisely the seroprevalence; therefore, it reports seroprevalence estimation. In conclusion, this study estimates the national serological prevalence of COVID-19 in Saudi Arabia to be 11%, with an apparent disparity between regions. This warrants better vaccination programs and vaccination coverage to increase the immuned population.

## Data Availability

All data are analysed and reported in the manuscript. Raw data are available by requesting it from the authors.

## Acknowledgment

We thank Dr. Ahmad Alaskar, KAIMRC Executive Director; Dr. Barrak Alsomaie, KAIMRC Operation Director; Dr. Azza Jadallah, Department of Pathology and Lab Medicine, KAMC; Dr. Ahmad Salman, University of Oxford; Dr. Ali Hajeer, Head of NGHA serology lab for their support.

## Author contribution

*Sample collection:* Areeb Albargawi, Abdullah G. Alharbi, Abdulaziz Alhazmi, Ali Alqarni, Ali Alfarhan, Fayhan Alroqi, Mohammed Bosaeed, Yaseen M. Arabi, Hosam M. Zowawi. *Sample testing:* Hind Albalawi, Mohammed Alenazi, Naif K. Alharbi. *Data analysis and Manuscript Writing:* Naif K. Alharbi, Omar Aldibasi, Suliman Alghnam, Anwar Hashem, Abdullah Algaissi.

## Supplementary file

**Table S1:** Demographics of blood donors and non-COVID-19 patients in sampled regions.

| Region | Population | Age Mean | Females No (%) | Males No (%) | Total number |
| --- | --- | --- | --- | --- | --- |
| Riyadh | Overall |  | 2025 | 1899 | 3924 |
| Riyadh | Blood Donors | 32.42 | 30 (2.52) | 1162 (97.48) | 1192 |
| Riyadh | Non-COVID-19 Patients | 42.40 | 1995 (73.02) | 737 (26.98) | 2732 |
| Jazan | Overall |  | 289 | 350 | 639 |
| Jazan | Blood Donors | 26.68 | 3 (3.75) | 77 (96.25) | 80 |
| Jazan | Non-COVID-19 Patients | 24.43 | 286 (51.16) | 273 (48.84) | 559 |
| Qassim | Overall |  | 724 | 2309 | 3033 |
| Qassim | Blood Donors | 29.77 | 46 (2.82) | 1587 (97.18) | 1633 |
| Qassim | Non-COVID-19 Patients | 50.91 | 678 (48.43) | 722 (51.57) | 1400 |
| Hail | Overall |  | 55 | 405 | 460 |
| Hail | Blood Donors | 30.33 | 0 (0.00) | 300 (100.00) | 300 |
| Hail | Non-COVID-19 Patients | 51.92 | 55 (34.38) | 105 (65.63) | 160 |
| Alahsa | Overall |  | 587 | 1291 | 1878 |
| Alahsa | Blood Donors | 32.13 | 32 (3.75) | 822 (96.25) | 854 |
| Alahsa | Non-COVID-19 Patients | 58.53 | 555 (54.20) | 469 (45.80) | 1024 |
| Makkah | Overall |  | 20 | 1012 | 1032 |
| Makkah City | Blood Donors | 32.28 | 5 (0.94) | 527 (99.06) | 532 |
| Jeddah City | Blood Donors | 32.59 | 15 (3.00) | 485 (97.00) | 500 |

**Table S2:** Regional population and COVID-19 infection rate in Saudi Arabia. Shaded table represents sampled regions.

| Region | Population size | COVID-19 incident rate (%) |
| --- | --- | --- |
| Saudi Arabia | 34,8 million | 1.00 |
| Riyadh | 8,014,678 | 0.88 |
| Jazan | 1,535,167 | 0.78 |
| Qassim | 1,389,929 | 0.92 |
| Hail | 685,423 | 0.98 |
| Alahsa | 4,787,375 | 1.8 |
| Makkah | 8,338,321 | 1 |
| Madinah | 2,083,326 | 1.27 |
| Tabuk | 891,813 | 0.53 |
| Northern border | 359,663 | 0.61 |
| Jawf | 498,081 | 0.22 |
| Baha | 466,946 | 0.81 |
| Asir | 2,166,983 | 1.24 |
| Najran | 569,875 | 1.08 |

## References

1. World Health Organisation. Coronavirus disease (COVID-19) pandemic [Internet]. 2020 [cited 2020 Nov 24]. Available from: https://www.who.int/emergencies/diseases/novel-coronavirus-2019

2. Young BE, Ong SWX, Ng LFP, Anderson DE, Chia WN, Chia PY, et al. Viral dynamics and immune correlates of COVID-19 disease severity. Clin Infect Dis an Off Publ Infect Dis Soc Am. 2020 Aug;

3. Lou B, Li T-D, Zheng S-F, Su Y-Y, Li Z-Y, Liu W, et al. Serology characteristics of SARS-CoV-2 infection after exposure and post-symptom onset. Eur Respir J [Internet]. 2020 Aug;56(2):2000763. Available from: http://erj.ersjournals.com/lookup/doi/10.1183/13993003.00763-2020

4. Guo L, Ren L, Yang S, Xiao M, Chang D, Yang F, et al. Profiling Early Humoral Response to Diagnose Novel Coronavirus Disease (COVID-19). Clin Infect Dis. 2020;

5. Dan JM, Mateus J, Kato Y, Hastie KM, Yu ED, Faliti CE, et al. Immunological memory to SARS-CoV-2 assessed for up to eight months after infection. bioRxiv [Internet]. 2020 Jan 1;2020.11.15.383323. Available from: http://biorxiv.org/content/early/2020/12/18/2020.11.15.383323.abstract

6. Wang K, Long Q-X, Deng H-J, Hu J, Gao Q-Z, Zhang G-J, et al. Longitudinal dynamics of the neutralizing antibody response to SARS-CoV-2 infection. Clin Infect Dis an Off Publ Infect Dis Soc Am. 2020 Aug;

7. Wang Y, Zhang L, Sang L, Ye F, Ruan S, Zhong B, et al. Kinetics of viral load and antibody response in relation to COVID-19 severity. J Clin Invest. 2020 Oct;130(10):5235–44.

8. Hashem AM, Algaissi A, Almahboub SA, Alfaleh MA, Abujamel TS, Alamri SS, et al. Early Humoral Response Correlates with Disease Severity and Outcomes in COVID-19 Patients. Viruses. 2020 Dec;12(12).

9. Long Q-X, Tang X-J, Shi Q-L, Li Q, Deng H-J, Yuan J, et al. Clinical and immunological assessment of asymptomatic SARS-CoV-2 infections. Nat Med. 2020 Aug;26(8):1200–4.

10. Poustchi H, Darvishian M, Mohammadi Z, Shayanrad A, Delavari A, Bahadorimonfared A, et al. SARS-CoV-2 antibody seroprevalence in the general population and high-risk occupational groups across 18 cities in Iran: a population-based cross-sectional study. Lancet Infect Dis. 2020 Dec;

11. Pollán M, Pérez-Gómez B, Pastor-Barriuso R, Oteo J, Hernán MA, Pérez-Olmeda M, et al. Prevalence of SARS-CoV-2 in Spain (ENE-COVID): a nationwide, population-based seroepidemiological study. Lancet (London, England). 2020 Aug;396(10250):535–44.

12. SeroTracker. SeroTracker [Internet]. [cited 2020 Oct 24]. Available from: https://serotracker.com/Explore

13. Rostami A, Sepidarkish M, Leeflang MMG, Riahi SM, Nourollahpour Shiadeh M, Esfandyari S, et al. SARS-CoV-2 seroprevalence worldwide: a systematic review and meta-analysis. Clin Microbiol Infect Off Publ Eur Soc Clin Microbiol Infect Dis. 2020 Oct;

14. Alserehi HA, Alqunaibet AM, Al-Tawfiq JA, Alharbi NK, Alshukairi AN, Alanazi KH, et al. Seroprevalence of SARS-CoV-2 (COVID-19) among healthcare workers in Saudi Arabia: comparing case and control hospitals. Diagn Microbiol Infect Dis. 2020 Nov;99(3):115273.

15. Interim guidance for use of pooling procedures in SARS-CoV-2 diagnostic, screening, and surveillance testing. August 1; 2020.

16. Evaluation of sample pooling for diagnosis of COVID-19 by real time-PCR: A resource-saving combat strategy. J Med Virol. 2020 Sep;

17. Saudi Ministry of Health. COVID 19 Dashboard: Saudi Arabia. 2021.

18. Mahallawi WH, Al-Zalabani AH. The seroprevalence of SARS-CoV-2 IgG antibodies among asymptomatic blood donors in Saudi Arabia. Saudi J Biol Sci. 2020;

19. Alandijany TA, El-Kafrawy SA, Al-Ghamdi AA, Qashqari FS, Faizo AA, Tolah AM, et al. Lack of Antibodies to SARS-CoV-2 among Blood Donors during COVID-19 Lockdown: A Study from Saudi Arabia. Healthcare. 2021 Jan;9(1):51.

20. Alharbi SA, Almutairi AZ, Jan AA, Alkhalify AM. Enzyme-Linked Immunosorbent Assay for the Detection of Severe Acute Respiratory Syndrome Coronavirus 2 (SARS-CoV-2) IgM/IgA and IgG Antibodies Among Healthcare Workers. Cureus. 2020 Sep;12(9):e10285.

2. Rijkers G, Murk J-L, Wintermans B, van Looy B, van den Berge M, Veenemans J, et al. Differences in antibody kinetics and functionality between severe and mild SARS-CoV-2 infections. J Infect Dis. 2020 Jul;

3. Young BE, Ong SWX, Ng LFP, Anderson DE, Chia WN, Chia PY, et al. Viral dynamics and immune correlates of COVID-19 disease severity. Clin Infect Dis an Off Publ Infect Dis Soc Am. 2020 Aug;

4. Lou B, Li T-D, Zheng S-F, Su Y-Y, Li Z-Y, Liu W, et al. Serology characteristics of SARS-CoV-2 infection after exposure and post-symptom onset. Eur Respir J. 2020 Aug;56(2):2000763.

5. Guo L, Ren L, Yang S, Xiao M, Chang D, Yang F, et al. Profiling Early Humoral Response to Diagnose Novel Coronavirus Disease (COVID-19). Clin Infect Dis. 2020;

13. Alserehi HA, Alqunaibet AM, Al-Tawfiq JA, Alharbi NK, Alshukairi AN, Alanazi KH, et al. Seroprevalence of SARS-CoV-2 (COVID-19) among healthcare workers in Saudi Arabia: comparing case and control hospitals. Diagn Microbiol Infect Dis. 2020 Nov;99(3):115273.

14. Interim guidance for use of pooling procedures in SARS-CoV-2 diagnostic, screening, and surveillance testing. August 1; 2020.

15. Evaluation of sample pooling for diagnosis of COVID-19 by real time-PCR: A resource-saving combat strategy. J Med Virol. 2020 Sep;

16. Saudi Ministry of Health. COVID 19 Dashboard: Saudi Arabia [Internet]. 2021 [cited 2020 Dec 31]. Available from: https://covid19.moh.gov.sa/

17. Mahallawi WH, Al-Zalabani AH. The seroprevalence of SARS-CoV-2 IgG antibodies among asymptomatic blood donors in Saudi Arabia. Saudi J Biol Sci. 2020;

18. Alandijany TA, El-Kafrawy SA, Al-Ghamdi AA, Qashqari FS, Faizo AA, Tolah AM, et al. Lack of Antibodies to SARS-CoV-2 among Blood Donors during COVID-19 Lockdown: A Study from Saudi Arabia. Healthcare. 2021 Jan 5;9(1):51.

19. Alharbi SA, Almutairi AZ, Jan AA, Alkhalify AM. Enzyme-Linked Immunosorbent Assay for the Detection of Severe Acute Respiratory Syndrome Coronavirus 2 (SARS-CoV-2) IgM/IgA and IgG Antibodies Among Healthcare Workers. Cureus. 2020 Sep;12(9):e10285.

